# Changes in healthcare workers’ knowledge, attitudes, practices, and stress during the COVID-19 pandemic

**DOI:** 10.1101/2021.01.19.21250126

**Authors:** Mohamad-Hani Temsah, Abdullah Al Huzaimi, Abdulkarim Alrabiaah, Nurah Alamro, Fahad Al-Sohime, Ayman Al-Eyadhy, Khalid Alhasan, Jameela A Kari, Ali Alhaboob, Amro Al Salmi, Wejdan AlMuhanna, Ibrahim Almaghlouth, Fadi Aljamaan, Rabih Halwani, Mazin Barry, Fahad Al-Zamil, Ahmad Al-Hadi, Sarah Al-Subaie, Amr Jamal, Ali Mohammed Somily

**Affiliations:** College of Medicine, King Saud University, Riyadh, Saudi Arabia; Department of Pediatrics, King Saud University Medical City, Riyadh, Saudi Arabia; Prince Abdullah Ben Khaled Coeliac Disease Chair, Faculty of Medicine, King Saud University, Riyadh, Saudi Arabia; Cardiac Science Department, King Saud University Medical City, Riyadh, Saudi Arabia; Department of Family and Community Medicine, King Saud University Medical City, Riyadh, Saudi Arabia; Prince Sattam bin Abdulaziz Research Chair for Epidemiology and Public Health, King Saud University, Riyadh, Saudi Arabia; Paediatric Nephrology Centre of excellence, Department of Paediatrics, King Abdulaziz University, Jeddah, Saudi Arabia; College of Medicine Research Center, King Saud University, Riyadh, Saudi Arabia; Adult Critical Care Department, King Saud University, King Saud University Medical City/King Khalid University Hospital, Riyadh, Saudi Arabia; Sharjah Institute of Medical Research, College of Medicine, University of Sharjah, Sharjah, United Arab Emirates; Infectious Disease Unit, Department of Internal Medicine, King Saud University, Riyadh, Saudi Arabia; Department of Psychiatry, College of Medicine, King Saud University and King Saud University Medical City, Riyadh, Saudi Arabia; SABIC Psychological Health Research and Applications Chair, College of Medicine, King Saud University, Riyadh, Saudi Arabia; Evidence-Based Health Care & Knowledge Translation Research Chair, King Saud University, Riyadh, Saudi Arabia; Department of Pathology and Laboratory Medicine, College of Medicine, King Saud University and King Saud University Medical City, Riyadh, Saudi Arabia

**Keywords:** COVID-19, changing KAP, healthcare worker

## Abstract

**Introduction:** Coronavirus disease 2019 (COVID-19) has caused an unprecedented health crisis around the world, not least because of its heterogeneous clinical presentation and course. The new information on the pandemic emerging daily has made it challenging for healthcare workers (HCWs) to stay current with the latest knowledge, which could influence their attitudes and practices during patient care.

**Methods:** This study is a follow-up evaluation of changes in HCWs’ knowledge, attitudes, and practices as well as anxiety levels regarding COVID-19 since the beginning of the pandemic. Data were collected through an anonymous, predesigned, self-administered questionnaire that was sent online to HCWs in Saudi Arabia.

**Results:** The questionnaire was sent to 1500 HCWs, with a 63.8% response rate (N=957). The majority of respondents were female (83%), and the most common age group was 31–40 years (52.2%). Nurses constituted 86.3% of the respondents. HCWs reported higher anxiety during the COVID-19 pandemic which increased from 4.91±2.84 to 8.6±2.27 on an 11-point Likert scale compared to other viral outbreaks. HCWs believed that their own preparedness as well as that of their hospital’s intensive care unit (ICU) or emergency room (ER) was higher during the COVID-19 pandemic than during the Middle East respiratory syndrome coronavirus pandemic (2012–2015). About 58% of HCWs attended one or more simulations concerning the management of COVID-19 patients in their ICU/ER, and nearly all had undergone N95 mask fit testing. The mean score of HCWs’ knowledge of COVID-19 was 9.89/12. For most respondents (94.6%), the perception of being at increased risk of infection was the main cause of anxiety related to COVID-19; the mean score of anxiety over COVID-19 increased from 4.91±2.84 before to 8.6±2.27 during the pandemic in Saudi Arabia.

**Conclusions:** HCWs’ anxiety levels regarding COVID-19 have increased since a pandemic was declared. It is vital that healthcare facilities provide more emotional and psychological support for all HCWs.

## Introduction

Coronavirus disease 2019 (COVID-19) has caused an unprecedented health crisis around the world and has surprised healthcare workers (HCWs) because of its wide dynamic and heterogeneous clinical presentation, from asymptomatic to critical illness leading to hospitalization and death.^[1]^ The rapid evolution of available literature and new information on the pandemic emerging daily has made it challenging for HCWs to stay current with the latest knowledge, which could influence their attitudes and practices during patient care. Additionally, basic principles of infection prevention and control (IPC) are more widely recognized and implemented by HCWs in Saudi Arabia, including hand hygiene, personal protective equipment (PPE) compliance, and N95 fit testing.^[2]^ The previous experience of Middle East respiratory syndrome coronavirus (MERS-CoV) outbreaks at several hospitals^[3-9]^ has enhanced IPC knowledge and practices among HCW, and commissioning bodies such as the Saudi Central Board for Accreditation of Healthcare Institutions have updated their definitions and standards for coronaviruses and acute respiratory syndromes to include COVID-19.^[10]^ As such, hospitals in Saudi Arabia were quick to adapt to and prepare for the pandemic before the first cases appeared in the country.^[11]^

In the present study, we assessed the anxiety levels and knowledge base of HCWs in a tertiary care hospital in Saudi Arabia during the pandemic as compared to its very beginning in order to determine the degree of preparedness of the healthcare system to not only to manage patients but also to meet the needs of HCWs, and identify potential deficiencies that can be improved by focused education and training.

## Methods

### Data collection

This was the second study for serial cross-sectional surveys among HCWs in Saudi Arabia during the COVID-19 pandemic. The survey was a pilot-validated, self-administered questionnaire sent online to HCWs that was adapted from our previously published study.^[12]^ We used multiple professional social media groups and email lists from our previous surveys during the MERS-CoV outbreak in 2014 and the early phase of the COVID-19 pandemic in early 2020.

Data were collected over 2 weeks (April 13–27, 2020). The first part of the survey addressed respondents’ characteristics (sex, age, etc.) and sources of information used during the pandemic.

We assessed the following domains for each participant: (1) knowledge concerning COVID-19; (2) attitudes toward IPC measures; and (3) changes in hygiene practices. Knowledge on COVID-19 was assessed using 6 questions consisting of true or false answers, with a score of 0–12 points assigned (Supplementary Table S1). The degree of change in HCWs’ attitudes was measured using a series of questions on a Likert-like scale from 1–5 (Supplementary Table S2). Hygiene practices were similarly measured to assess the degree of behavioral change in HCWs on scale of 1–4 (Supplementary Table S3).

HCW anxiety and preparedness with respect to COVID-19 were evaluated based on self-reported perceived anxiety over COVID-19 vs MERS-CoV vs seasonal influenza (11-point Likert scale), along with the level of worry over COVID-19 infection in the previous 2 weeks. HCWs’ perceptions of their own preparedness as well as that of their hospital with respect to the COVID-19 pandemic vs the previous MERS-CoV outbreak were compared using an 11-point Likert scale. Additionally, the availability of a psychological support system, HCWs’ adherence to seasonal influenza vaccine, and participation in COVID-19 simulation training, N95 mask fit testing and sources of anxiety for each HCWs were evaluated. Data collected from HCWs through the online survey were entered into a secure spreadsheet in SPSS (SPSS Inc, Chicago, IL, USA). All data was treated with strict confidentiality, with the anonymity of respondents maintained throughout the study.

### Statistical analysis

Descriptive statistics approaches with mean, median, and standard deviation were applied to continuous variables, while percentages were used for dichotomous variables. The 2-sample t test was used to evaluate continuous scores and the Z-test was used to compare proportions. The multiple response dichotomy analysis was applied to describe the healthcare workers sources of covid19 Information’s.

A multivariate logistic regression model was used to explore the associations between the outcome variable the level of anxiety from COVID-19 and demographic characteristics of HCWs, attitude and hygiene practice scores, and sources of anxiety, the associations between predictors and the outcome was expressed as Odds Ratio and 95% Confidence Interval. The SPSS IBM Version21 was used for the data analysis, the excel program was used for creating figures and depictions, the p-value statistical significance was considered at 0.050 level. The Institutional Review Board of King Saud University Medical City approved the study. HCWs gave informed consent prior to participation.

## Results

Of the 1500 HCWs who were contacted, 1453 agreed to participate in the study. Participants with information missing on more than 50% of the questionnaire were excluded.

### Demographic and sources of information data

Data from the questionnaire were completed by 957 (63.8%) HCWs and were included in the final analysis. Most respondents were employed at public governmental tertiary hospitals in Riyadh, Saudi Arabia; the majority were female (83%) and between 31–40 years of age (52.2%). Nurses constituted 86.3% of the study population. Most respondents worked in general hospital wards (26.5%), followed by outpatient clinics (18.8%) and adult emergency rooms (ERs) (14.7%) (Table 1). HCWs obtained information on COVID-19 from multiple sources (Figure 1).

**Table 1.** Sociodemographic and professional characteristics of healthcare workers (N=957)

|  | Frequency | Percentage |
| --- | --- | --- |
| **Sex** |  |  |
| Male | 163 | 17 |
| Female | 794 | 83 |
| Age (years), mean (SD) |  | 36.69 (7.79) |
| 21–30 years | 192 | 20.1 |
| 31–40 years | 500 | 52.2 |
| 41–50 years | 206 | 21.5 |
| ≥51 years | 59 | 6.2 |
| **Clinical role** |  |  |
| Consultant | 69 | 7.2 |
| Assistant consultant | 22 | 2.3 |
| Registrar | 13 | 1.4 |
| Resident | 27 | 2.8 |
| Nurse | 826 | 86.3 |
| **Department** |  |  |
| Adult ICU | 139 | 14.5 |
| Pediatric ICU | 75 | 7.8 |
| Adult ER | 141 | 14.7 |
| Pediatric ER | 51 | 5.3 |
| Isolation ward | 34 | 3.6 |
| General ward | 254 | 26.5 |
| OPD | 180 | 18.8 |
| Operation room | 40 | 4.2 |
| Other units (dialysis, radiology, other services) | 43 | 4.5 |
| **Hospital sector** |  |  |
| Private | 32 | 3.3 |
| Public/governmental | 925 | 96.7 |
| **Hospital setting** |  |  |
| Tertiary | 850 | 88.8 |
| Non-tertiary (secondary etc) | 107 | 11.2 |
| Hospital location |  |  |
| Riyadh | 907 | 94.8 |
| Other city | 50 | 5.2 |

ER, emergency room; ICU, intensive care unit; OPD, outpatient department; SD, standard deviation.

**Figure 1.**
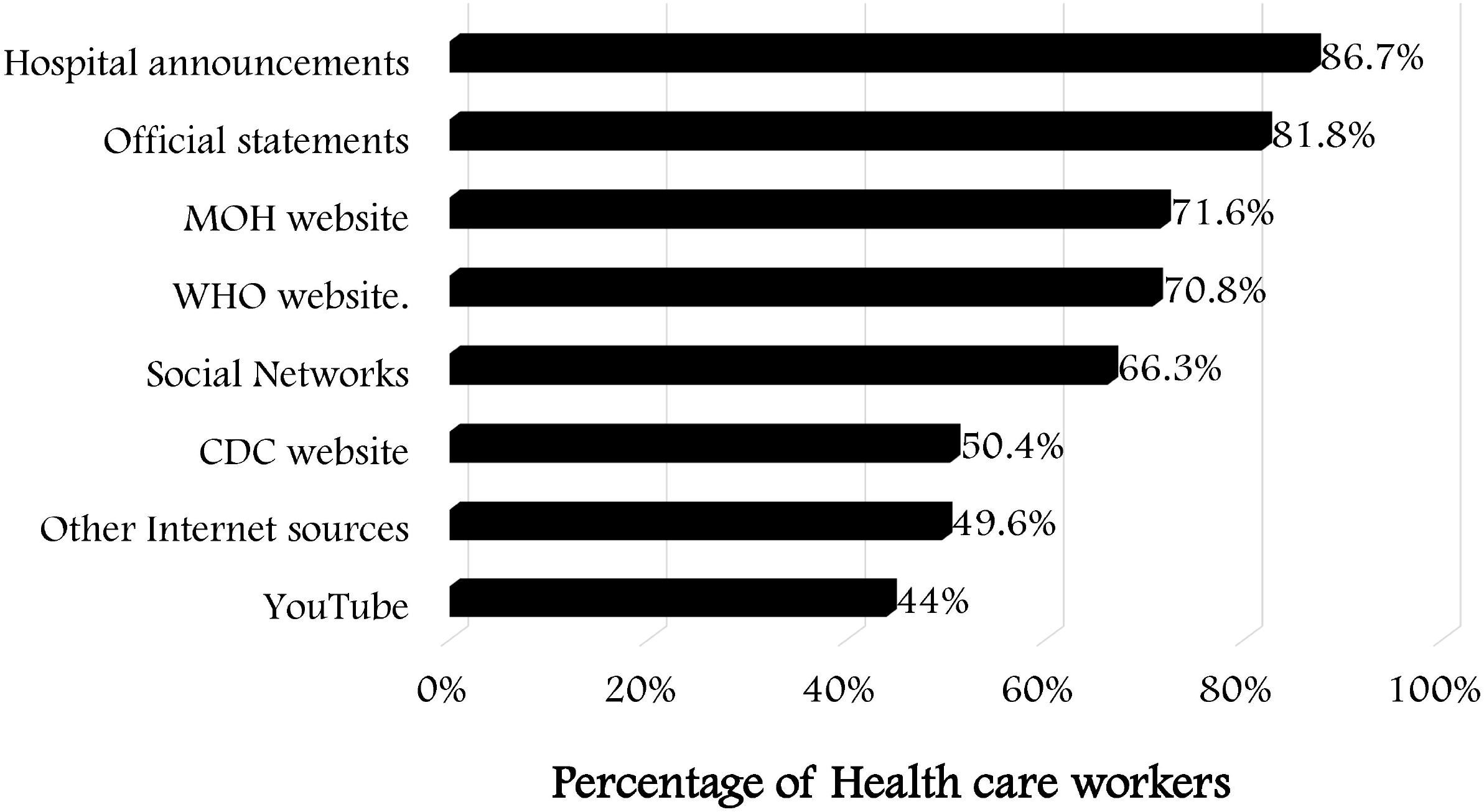
Sources of information regarding COVID-19 used by HCWs.

### HCW anxiety and preparedness with respect to COVID-19 during the pandemic

HCWs’ self-reported anxiety levels regarding COVID-19, MERS-CoV, and seasonal influenza were compared (Figure 2). The highest levels of anxiety were experienced during the COVID-19 pandemic compared to other viral outbreaks. HCWs believed that their own preparedness and those of their hospital’s ER or intensive care unit (ICU) was higher during the COVID-19 pandemic than during the 2012–2015 MERS-CoV outbreaks (Table 2); however, 88% felt more worried about the former.

**Table 2.** Descriptive analysis of healthcare workers’ perceived stress, anxiety, and preparedness practices during the COVID-19 pandemic

|  | Frequency (%) /<br>Scale mean (SD) |
| --- | --- |
| **Anxiety** |  |
| **A) Rate the anxiety you have experienced during the following epidemic/pandemics: (11-point Likert scale)** |  |
| Current COVID-19 outbreak/pandemic | 8.56 (2.27) |
| MERS-CoV | 6.40 (2.35) |
| Seasonal flu | 4.94 (2.65) |
| **B) How worried were you in the past week about contracting COVID-19? (5-point Likert scale)** | 3.44 (1.10) |
| **C) Compared to the MERS-CoV epidemic in 2012–2015, how do you feel about the recent COVID-19 (2019-NCOV) pandemic?** |  |
| Same level of stress during both pandemics | 73 (7.6%) |
| I have felt more stressed during the COVID-19 (2019-NCOV) pandemic | 843(88.1%) |
| I felt more stressed during the previous MERS-CoV epidemic | 41(4.3%) |
| **Preparedness** |  |
| Your preparedness for the COVID-19 pandemic (11-point Likert scale) | 8.84 (2.03) |
| Your preparedness for the previous MERS-CoV outbreak (11-point Likert scale) | 8.53 (2.35) |
| Your ICU/ER department's preparedness during the COVID-19 pandemic (11-point Likert scale) | 9.10 (2.02) |
| Your ICU/ER department's preparedness during the MERS-CoV epidemic (11-point Likert scale) | 8.83 (2.19) |
| **Have you received the influenza vaccine (flu shot) this season?** |  |
| Yes, I receive it every year | 0757 (79.1%) |
| Yes, only this year | 50 (5.2%) |
| No | 150(15.7%) |
| **Did you undergo an N95 mask fit test?** |  |
| Yes, in 2020 | 471 (49.2%) |
| Yes, before this year | 463 (48.4%) |
| Never | 23 (2.4%) |
| **Did you have a simulation about possible COVID-19 patients in your hospital's ICU/ER?** |  |
| Yes, once | 252 (26.3%) |
| Yes, several simulation scenarios | 306 (32%) |
| No | 399 (41.7%) |
| **Does your hospital have psychological support services to support HCWs during COVID-19?** |  |
| Yes | 709 (74.1%) |
| No | 248 (25.9%) |

2019-NCOV, 2019 novel coronavirus; COVID-19, coronavirus disease 2019; ER, emergency room; HCW, healthcare worker; ICU, intensive care unit; MERS-CoV, Middle East respiratory syndrome coronavirus; SD, standard deviation.

**Figure 2.**
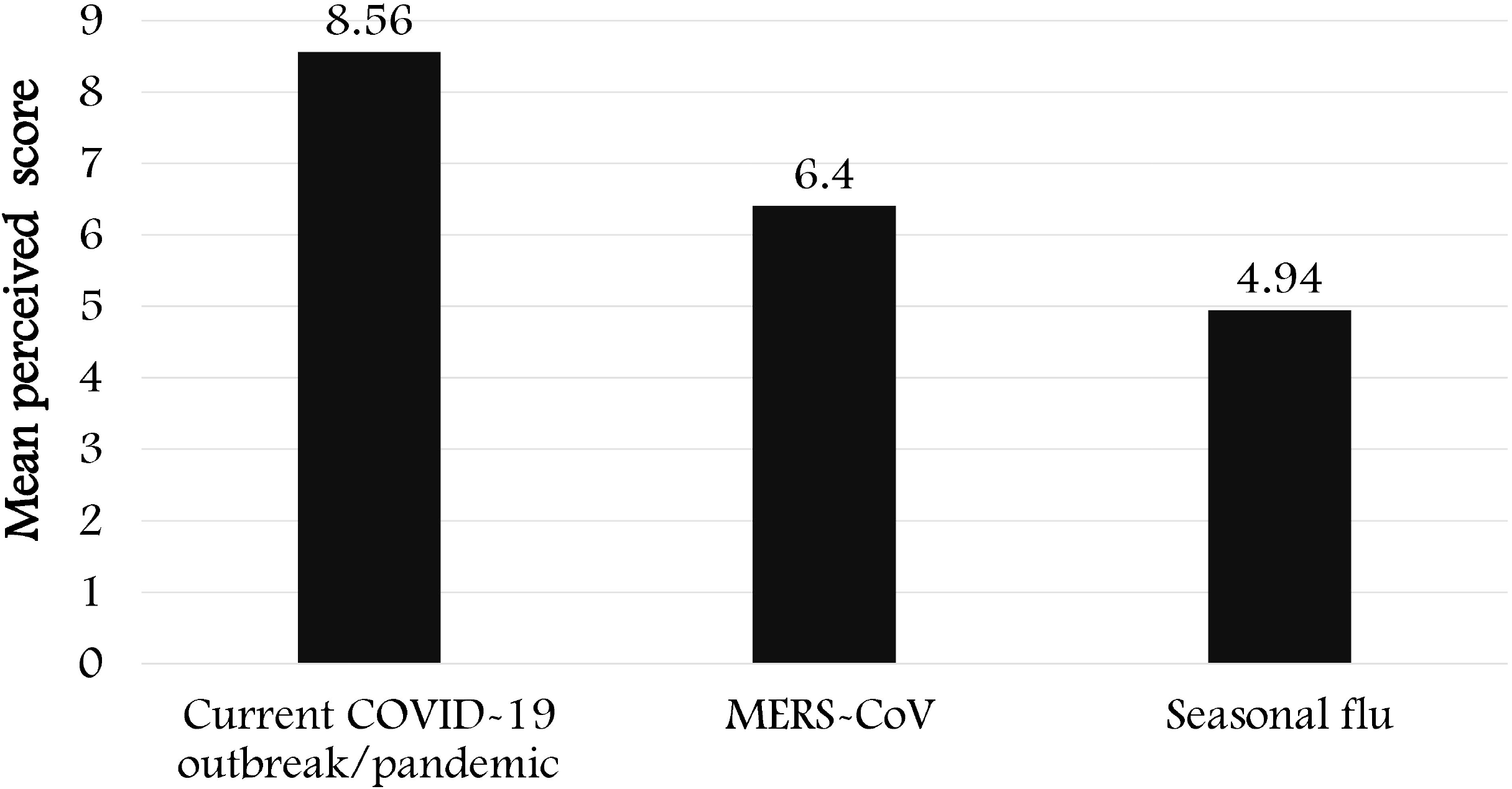
HCWs’ self-rated mean anxiety related to various viral outbreaks (11-point Likert-like scale).

About 97% of respondents had undergone N95 mask fit testing in 2020 or in the previous year, but only 58% had attended at least one simulation session related to managing COVID-19 patients in their hospital’s ICU/ER. About one-quarter of HCWs worked at hospitals where no psychological support system was available during the COVID-19 pandemic.

### Knowledge, attitudes, and practices (KAP) of HCWs with respect to COVID-19

The mean score for HCWs’ knowledge pertaining to COVID-19 was 9.89 out of 12. HCWs’ attitude toward infection control measures were measured on a 5-point Likert-like scale, with a mean score of 4.42±1.1 (Table 3), which indicated that most participants agreed or strongly agreed with level of importance of implementing their hospital’s IPC measures (Supplementary Table S2).

**Table 3.** Descriptive statistics of HCWs’ perceptions of COVID-19.

|  | **Mean (SD)** |
| --- | --- |
| **Knowledge of COVID-19 (0 to 12 points)** | 9.89 (1.98) |
| **Attitudes toward infection control measures (level of agreement, 5-point Likert scale)** | 4.42 (1.1) |
| **Hygiene practice improvement score (level of behavior change, 4-point Likert scale)** | 3.82 (0.32) |

COVID-19, coronavirus disease 2019; SD, standard deviation.

The mean hygiene practice score was 3.8±0.32, representing a moderate-to-large change in self-rated compliance with hygiene practices and behaviors (e.g., hand hygiene) (Supplementary Table S3).

### Sources of anxiety among HCWs

Sources of perceived anxiety related to COVID-19 in HCWs are shown in Figure 3. Most HCWs (94.6%) were concerned about the risk of acquiring COVID-19 during the pandemic, while more than half of the group was worried about the depletion of PPE at their hospital in general or in specifically high-risk departments such as the ICU or ER. On the other hand, the HCWs described several coping interventions to handle the stress during the pandemic, as demonstrated in the “word cloud” representation in Figure 4.

**Figure 3.**
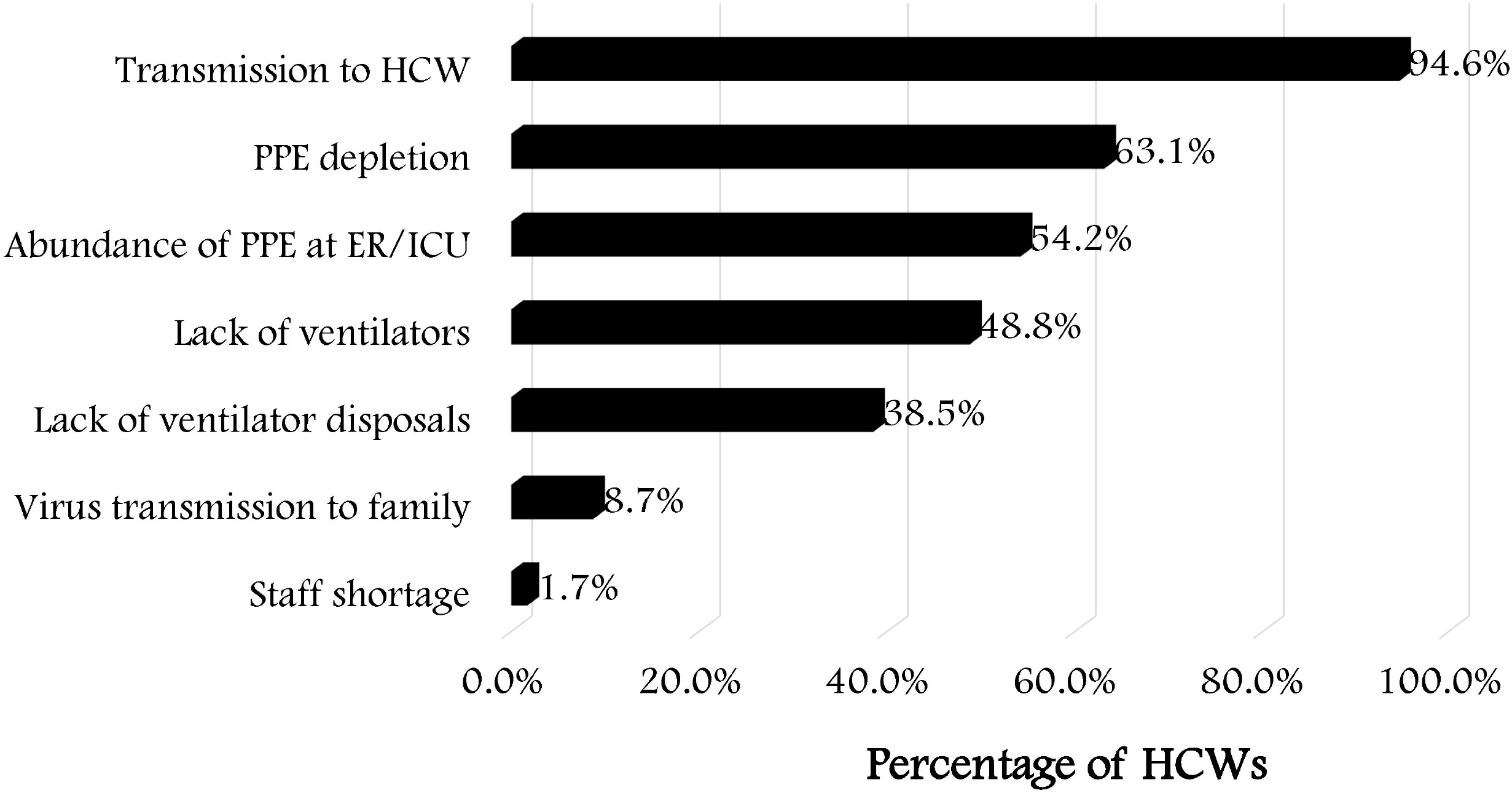
Sources of concern and anxiety related to COVID-19 among HCWs.

**Figure 4.**
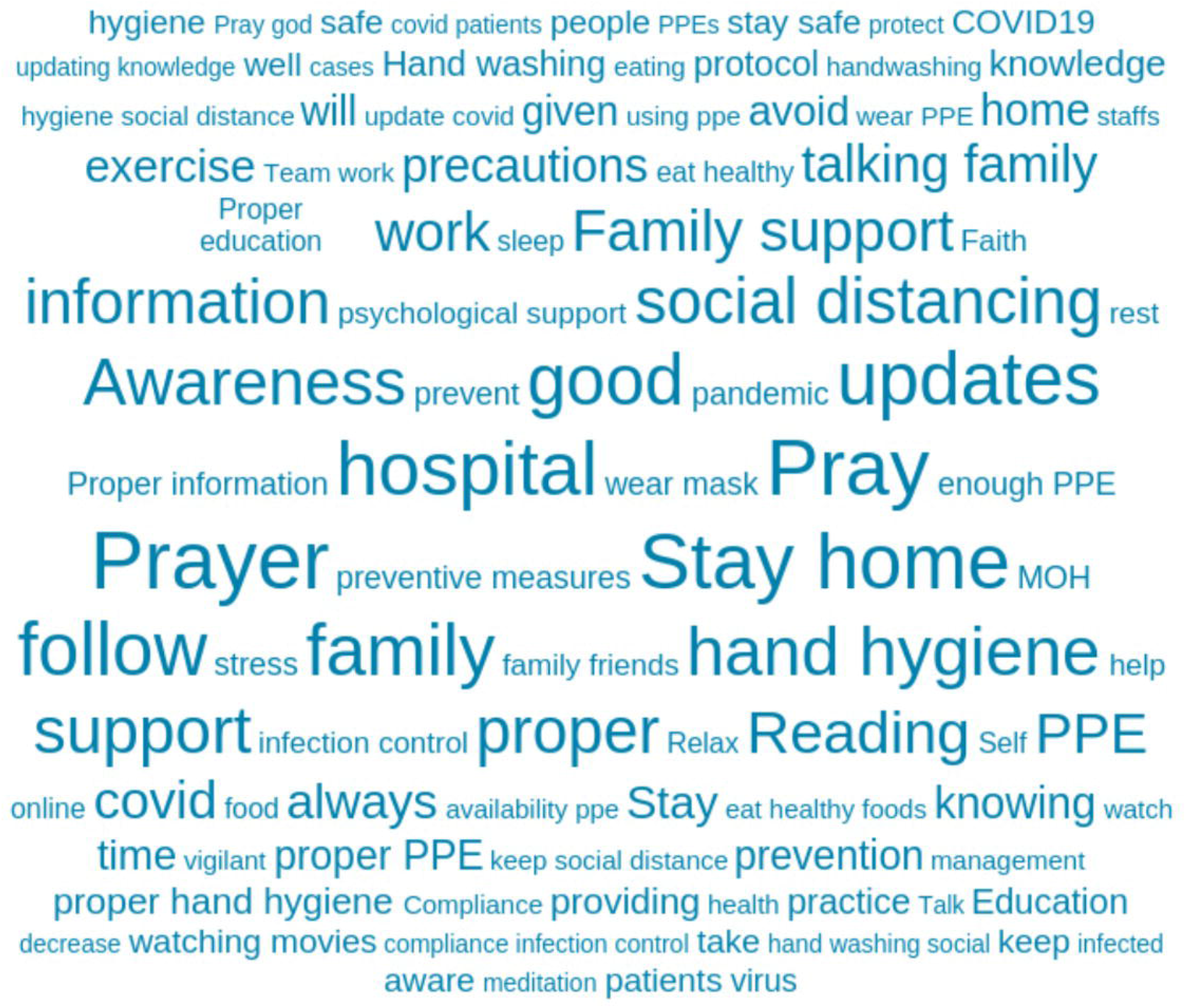
Word cloud representation of what best made healthcare workers cope with the stress during the pandemic (N = 957)

### Factors associated with HCWs’ perceived anxiety with respect to COVID-19

Factors contributing to HCWs’ perceived anxiety over COVID-19 during the pandemic were analyzed using a multivariate logistic regression model (Table 4). The model was significant overall (f[17,939]=5.98, P<0.001).

**Table 4.**
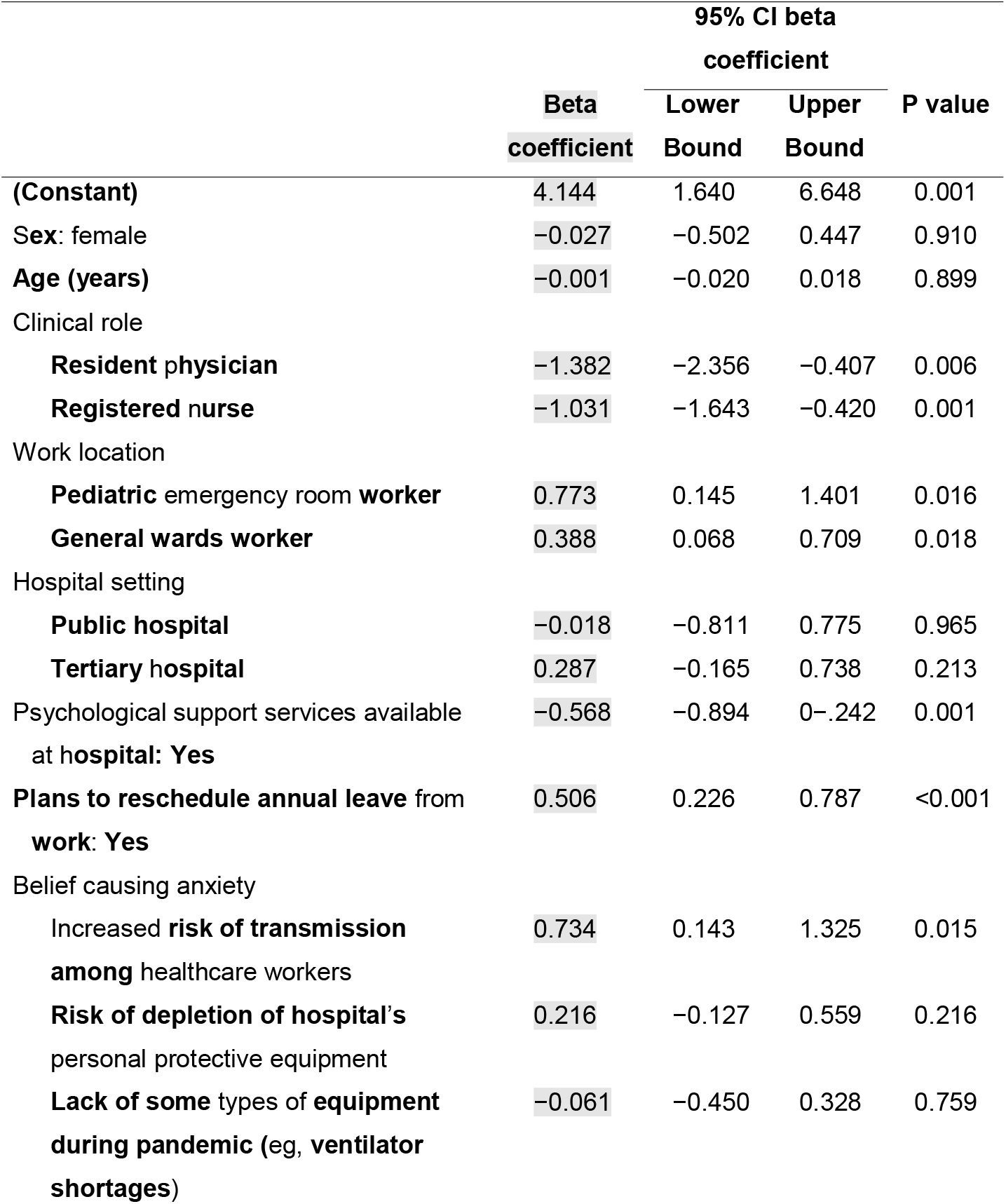
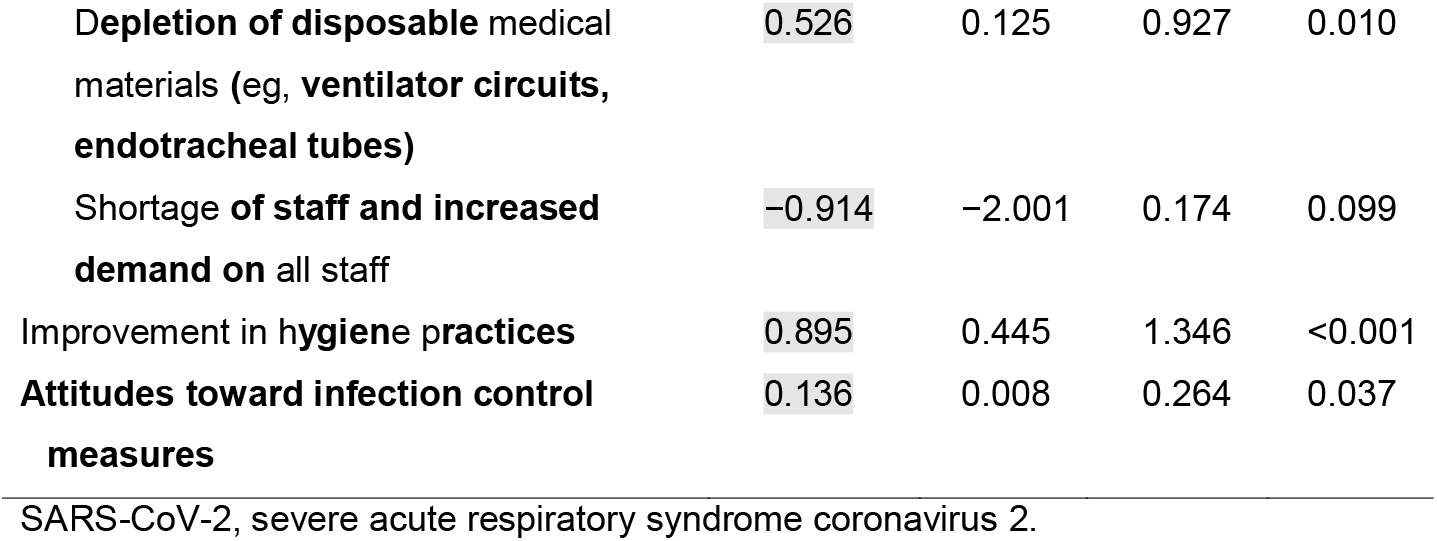
Multivariate linear regression analysis of healthcare workers’ perceived anxiety caused by COVID-19 (N=956)

|  | Beta<br>coefficient | 95% CI beta<br>coefficient |  | P value |
| --- | --- | --- | --- | --- |
|  |  | Lower<br>Bound | Upper<br>Bound |  |
| **(Constant)** | 4.144 | 1.640 | 6.648 | 0.001 |
| Sex: female | -0.027 | -0.502 | 0.447 | 0.910 |
| **Age (years)** | -0.001 | -0.020 | 0.018 | 0.899 |
| Clinical role |  |  |  |  |
| **Resident physician** | -1.382 | -2.356 | -0.407 | 0.006 |
| **Registered nurse** | -1.031 | -1.643 | -0.420 | 0.001 |
| Work location |  |  |  |  |
| **Pediatric emergency room worker** | 0.773 | 0.145 | 1.401 | 0.016 |
| **General wards worker** | 0.388 | 0.068 | 0.709 | 0.018 |
| Hospital setting |  |  |  |  |
| **Public hospital** | -0.018 | -0.811 | 0.775 | 0.965 |
| **Tertiary hospital** | 0.287 | -0.165 | 0.738 | 0.213 |
| Psychological support services available<br>at hospital: **Yes** | -0.568 | -0.894 | 0-.242 | 0.001 |
| **Plans to reschedule annual leave from**<br>**work: Yes** | 0.506 | 0.226 | 0.787 | <0.001 |
| Belief causing anxiety |  |  |  |  |
| Increased **risk of transmission**<br>**among** healthcare workers | 0.734 | 0.143 | 1.325 | 0.015 |
| **Risk of depletion of hospital's**<br>**personal protective equipment** | 0.216 | -0.127 | 0.559 | 0.216 |
| **Lack of some types of equipment**<br>**during pandemic (eg, ventilator**<br>**shortages)** | -0.061 | -0.450 | 0.328 | 0.759 |
| **Depletion of disposable** medical materials (eg, **ventilator circuits, endotracheal tubes**) | 0.526 | 0.125 | 0.927 | 0.010 |
| **Shortage of staff and increased demand on** all staff | -0.914 | -2.001 | 0.174 | 0.099 |
| Improvement in **hygiene practices** | 0.895 | 0.445 | 1.346 | <0.001 |
| **Attitudes toward infection control measures** | 0.136 | 0.008 | 0.264 | 0.037 |

SARS-CoV-2, severe acute respiratory syndrome coronavirus 2.

Neither sex nor age was correlated with perceived anxiety over COVID-19. However, the clinical role and work location of the HCW showed positive associations with perceived anxiety level during the pandemic: resident physicians and nurses had lower mean perceived anxiety than HCWs in other roles (e.g. consultant or assistant consultant) (both P<0.010). Additionally, HCWs at general hospitals (including in COVID-19 wards) and pediatric ER had higher mean perceived anxiety than their colleagues in other departments (P=0.018 and 0.016, respectively).

Hospital setting and sector were not correlated with mean perceived anxiety of HCWs during the COVID-19 pandemic; however, HCWs at hospitals equipped with a psychological support system dedicated to staff during the pandemic reported a lower mean perceived anxiety than those working in hospitals without such a system (P=0.001).

HCWs who believed that the medical staff were at higher risk of contracting COVID-19 and for whom the possibility of shortage of medical supplies and disposable materials (e.g., ventilator circuits) were a source of anxiety had a significantly higher mean perceived anxiety level than other HCWs (P=0.015 and 0.010, respectively), although potential shortages in PPEs or equipment such as ventilators were not a significant source of anxiety. In addition, the fear of staff shortage during the pandemic did not significantly affect HCWs’ mean perceived anxiety. HCWs’ improvement in hygiene practice score was positively and significantly correlated with mean perceived anxiety over COVID-19 (P<0.001); additionally, their attitude toward hygiene practices was associated with a significantly higher mean anxiety (P=0.037)

Additional Bivariate analysis were done and revealed a significantly higher mean anxiety level over COVID-19 in HCWs at hospitals without support systems than in those at hospitals where such systems were available (9.02±1.88 vs 8.46±2.37, P<0.001) consistent with multivariate linear regression analysis. We also found that HCWs who used social networks as a source of information had a higher mean anxiety over COVID-19 than those who did not use social media (8.72±2.23 vs 8.39±2.33, P=0.032) (Table S4).

### HCW anxiety and KAP before vs during the COVID-19 pandemic

We compared the results of the present study (Phase II), which was carried out during the active COVID-19 pandemic in Saudi Arabia (April 13–27, 2020), with those of our previous study (Phase I; February 5–16, 2020), which was conducted by the same research team immediately before the first case of COVID-19 was reported in Saudi Arabia.

HCWs’ mean scores of perceived anxiety over COVID-19 increased from 4.91±2.84 to 8.6±2.27 on an 11-point Likert scale (Table 5). The degree of anxiety over other viral outbreaks, including MERS-CoV and seasonal influenza, similarly increased, as did concern over contracting COVID-19.

**Table 5.**
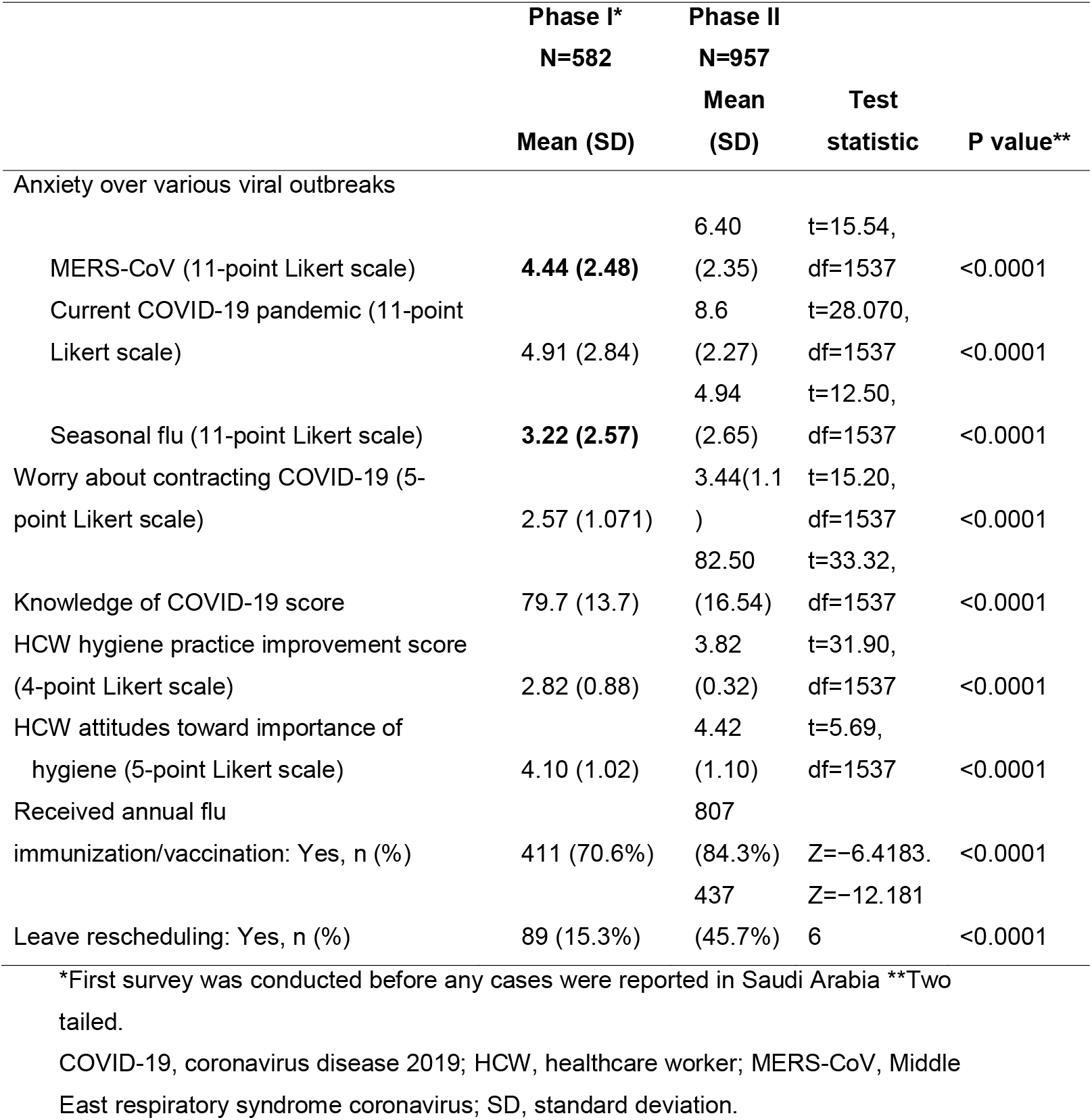
Bivariate comparisons with 2-sample mean and proportion differences in healthcare workers’ perceptions and behaviors before and during the COVID-19 pandemic

The Knowledge of COVID-19 in the second survey exceeded that measured in February 2020 (t=33.32, df=1537, p<0.0001 (2-sample t test), which is expected based on the elapsed time and exposure of the respondents to COVID-19 within that time frame. Likewise, scores for HCWs’ hygiene practices and attitudes toward hygiene increased significantly in the current survey compared to those measured in February (P<0.001).

The proportion of HCWs who received the annual influenza vaccine this year increased significantly during the pandemic as compared to before (Z=-6.4183, P<0.0001). The proportion of HCWs who intended to reschedule their annual leave also increased from 15.3% in February to 45.7% in April 2020 (Z=-12.1816, P<0.001).

## Discussion

The participants were predominantly female (83%) and most were nurses (86.3%), which is comparable to the demographic profile of the previous studies conducted in early 2020 prior to the COVID-19 outbreak in Saudi Arabia.^[13–16]^ Most respondents were between the ages of 31 and 40 years; some studies have shown that medical staff aged 31–40 years were more worried about becoming infected with severe acute respiratory syndrome coronavirus 2 (SARS-CoV-2, the causative agent of COVID-19) and infecting their families compared to other age groups.^[13, 15]^ A large proportion of the study participants were from acute care areas (42.42%), which is similar to other published work.

The current COVID-19 pandemic has been accompanied by an overabundance of information that has made it difficult to obtain accurate facts as most individuals rely on social media for this purpose;^[6]^ this contributes to higher levels of anxiety.^[17]^ Classic forms of media (i.e., television and newspapers) as well as social media affect the attitudes of both the public and health professionals, while consulting reliable sources of information is a basic condition for IPC. Compared to before the COVID 19 pandemic, hospital announcements were the top source of information used by HCWs during the pandemic (77.8% vs 86.7%), as was the case for the MERS-CoV outbreak.^[12]^ This finding highlights the importance of having a dedicated team to provide accurate information from trusted sources. News on social networks was a source of information for 61.7% of HCWs before and 66.3% during the pandemic, which may have contributed to greater anxiety in our study participants as bivariate analysis showed a higher mean anxiety over COVID-19 found that HCWs who used social networks as a source of information, although the use of unofficial and nonscientific sources of data was discouraged in awareness campaigns conducted by hospitals. Use of the Ministry of Health and World Health Organization websites as a source of information by HCWs increased from 42.3% to 71.6% and from 50% to 70.8%, respectively, between the 2 surveys.

Healthcare simulation tests have been useful for managing the COVID-19 pandemic. However, there are also practical constraints on running in situ simulations during a pandemic, such as the need for physical distancing, rigorous sanitization measures for mannequins and training equipment, and heightened anxiety among simulation participants; indeed, the fear of acquiring the infection could explain why 41.7% did not attend the simulation tests.^[18]^ In contrast, 97.65% of HCWs underwent N95 mask fit testing; this high percentage compared to the simulation test may be explained by the fact that the former was performed individually and therefore elicited less anxiety in HCWs in terms of the risk of acquiring COVID-19, and that such testing has been mandated for all HCW.

Surveys are commonly used to identify knowledge gaps and behavioral patterns in order to implement effective measures for improvement of processes and practices. We observed that knowledge scores were higher among participants in Phase II (during the COVID-19 pandemic) than in Phase I (pre-pandemic); the trend is similar to that observed during the previous MERS-CoV outbreak.^[12, 19]^ Most study participants had a high level of knowledge concerning SARS-CoV-2 infection and modes of transmission; this is expected given their profession as healthcare workers. However, previously published data from a KAP survey in the general population in China during the COVID- 19 pandemic revealed a high rate of correct responses in the knowledge questionnaire, which the authors attributed to the high educational level of the participants and the intensified public health education programs.^[20]^ Other studies have reported variable findings regarding KAP among HCWs during Ebola and Zika virus outbreaks. For example, satisfactory knowledge of Ebola virus disease without corresponding good practices was reported among Nigerian HCWs.^[21]^ This increase in HCWs knowledge could improve perceptions and positive attitudes that can translate into good practices, aiding in the prevention and management of infectious diseases. In our previous phase 1 study, the knowledge score was significantly associated with positive attitude and practice scores: that is, HCWs with a high level of knowledge had more positive attitudes and perceptions toward preventive measures and engaged to a greater extent in IPC practices. Others have reported similar associations in KAP surveys of COVID- 19 and other infectious diseases.^[22-24]^

The HCWs in our study reported a high level of anxiety with respect to COVID- 19, with 88% having more anxiety as compared to the previous MERS-CoV outbreak. Similarly, both local and international studies have found that anxiety levels were high among HCWs during both pandemics.^[25-27]^ A recent systematic review of 115 publications concluded that all coronavirus outbreaks (SARS, MERS-CoV, and COVID- 19) had a substantial impact on the physical and mental health of HCWs.^[27]^

The availability of hospital-based psychological support was associated with decreased levels of anxiety among HCWs in our survey. Psychological support is critical for the well-being of frontline HCWs.^[28]^ However, establishing psychological support services in Hospitals might not be enough to guarantee their efficacy, for instance in the case where there are obstacles to their access by HCWs who need them.^[29]^ One study proposed specific measures that healthcare managers should implement to protect the mental health of healthcare staff: managers must be honest about current and future situations and have regular meetings to discuss protocols and HCWs’ wellbeing; and after the crisis has passed, they must actively monitor, support, and—where necessary—provide treatment to HCWs.^[30]^ In accordance with this issue, the United Nations issued a policy report on the importance of HCWs’ mental health, and encouraged all parties to facilitate HCWs’ access to mental health services.^[31]^

Risk of being infected with SARS-CoV-2 was the highest source of anxiety among HCWs in our study, followed by depletion of PPE at their hospital in general or more specifically in the ICU/ER departments. These findings are similar to those of a study of 69 HCWs who participated in multiple listening sessions. There were 7 sources of anxiety, ranging from fear of not having enough equipment and of contracting COVID-19 or transmitting it to their loved ones, to fear of not performing well when they were needed in areas beyond their expertise.^[32]^ Such anxiety could affect the confidence of HCWs in themselves as well as general trust in the healthcare system. In China, HCWs who were interviewed reported that they did not require psychological help but needed enough protective supplies and more uninterrupted rest time.^[29]^ Another study examining factors related to HCWs’ psychological difficulties found that infection of colleagues and family members, protective measures, and medical violence were among the main concerns of HCWs in areas affected by COVID-19.^[33-34]^

This study had some limitations. Recall bias may have affected some participants’ responses, and most participants were nurses and females. Another issue is that many HCWs in our cohort had experienced the previous MERS-CoV epidemic; therefore, additional research in other settings are needed in order to determine whether our findings are generalizable.

## Conclusion

HCWs’ anxiety levels over COVID-19 increased after the pandemic was declared. Healthcare facilities need to provide more emotional and psychological support for all HCWs, including psychological first aid delivered through webinars to each unit on topics such as dealing with anxiety and insomnia, peer support, practicing self-care, and support for moral distress; as well as providing individual support sessions.

## Data Availability

There is no data in this paper. It is purely a modeling study.

## Ethics approval and consent to participate

Institutional Review Board, King Saud University, IRB # 20/0064/IRB

## Consent for publication

Granted upon manuscript acceptance

## Competing interests

The authors of this work have nothing to disclose.

## Funding

The authors are grateful to the Deanship of Scientific Research, King Saud University for funding through Vice Deanship of Scientific Research Chairs.

---

**Table S1.** Healthcare workers’ knowledge on COVID-19.

|  | **Incorrectly**<br>**answered, n**<br>**(%)** | **Correct**<br>**answered, n**<br>**(%)** |
| --- | --- | --- |
| COVID-19 can be transmitted by contacting patients'<br>stools and then touching the nose or eyes (T) | 46 (4.8) | 911 (95.2) |
| COVID-19 can be transmitted through contact with<br>packages shipped from China (F) | 425 (44.4) | 532 (55.6) |
| COVID-19 causes severe respiratory symptoms (T) | 17 (1.8) | 940 (98.2) |
| COVID-19 causes severe neurologic symptoms (F) | 419 (43.8) | 538 (56.2) |
| There is no vaccine against SARS-CoV-2 virus<br>infection (T) | 41 (4.3) | 916 (95.7) |
| There is no specific treatment for COVID-19 (T) | 57 (6) | 900 (94) |
| COVID-19, coronavirus disease 2019; F, false; SARS-CoV-2, severe acute respiratory<br>syndrome coronavirus 2; T, true |  |  |

**Table S2.** Healthcare workers’ attitudes toward SARS-CoV-2 infection control measures.

| **Attitude toward domestic hygienic practices** | **Level of importance\*,**<br>**mean (SD)** |
| --- | --- |
| It is important to wear face masks at work to prevent SARS-CoV-2 infection | 4.38 (1.22) |
| It is important to maintain good indoor ventilation to the spread of COVID-19 | 4.22 (1.23) |
| It is important to avoid going to crowded places during flu season | 4.52 (1.17) |
| Hand hygiene is very important to protect against SARS-CoV-2 infection | 4.57 (1.18) |

\*Likert scale score of 1 to 5 (1, strongly disagree; 2, disagree; 3, neutral; 4, agree; 5, strongly agree).
COVID-19, coronavirus disease 2019; SARS-CoV-2, severe acute respiratory syndrome coronavirus 2; SD, standard deviation

**Table S3.**
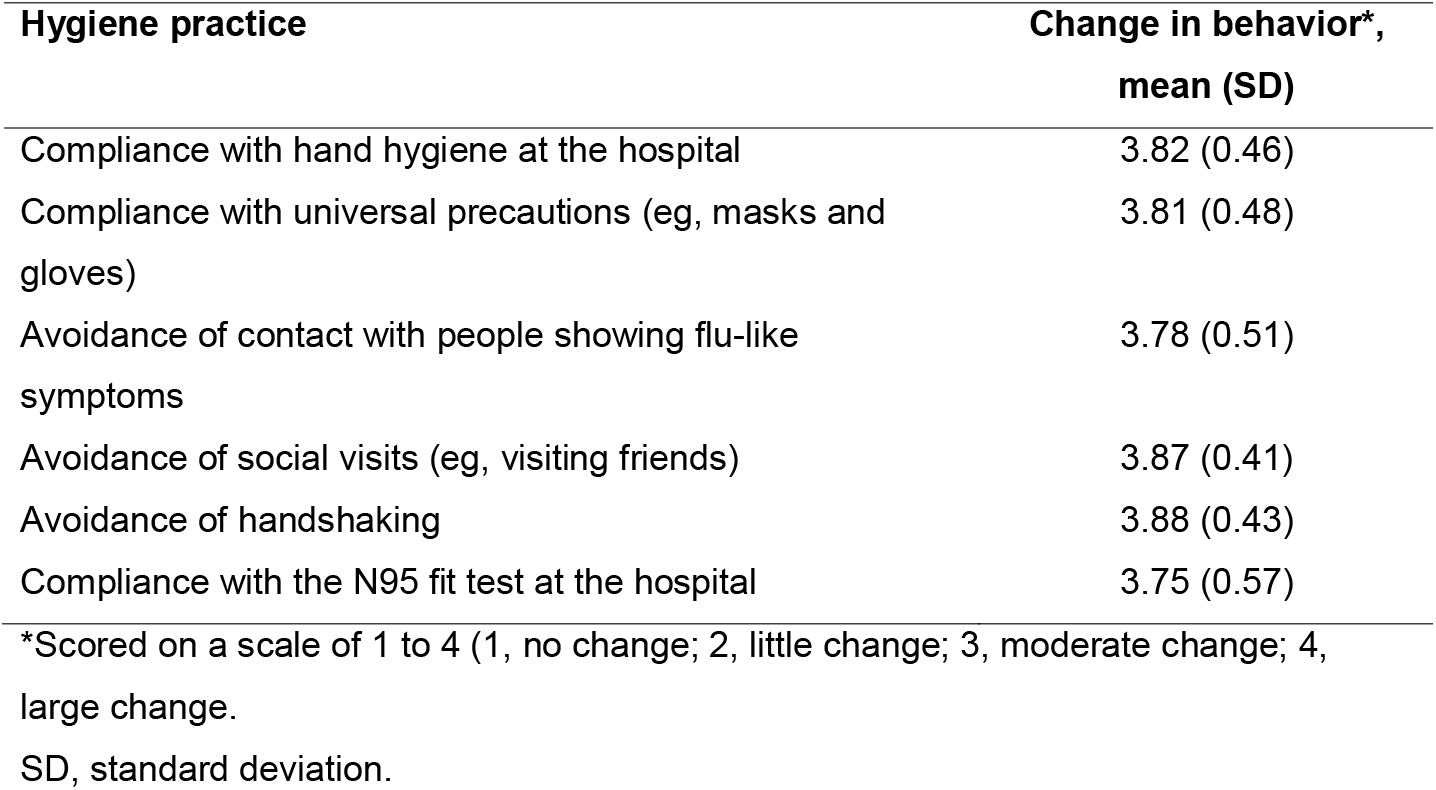
Changes in healthcare workers’ hygiene practices.

| **Hygiene practice** | **Change in behavior\*,**<br>**mean (SD)** |
| --- | --- |
| Compliance with hand hygiene at the hospital | 3.82 (0.46) |
| Compliance with universal precautions (eg, masks and gloves) | 3.81 (0.48) |
| Avoidance of contact with people showing flu-like symptoms | 3.78 (0.51) |
| Avoidance of social visits (eg, visiting friends) | 3.87 (0.41) |
| Avoidance of handshaking | 3.88 (0.43) |
| Compliance with the N95 fit test at the hospital | 3.75 (0.57) |

\*Scored on a scale of 1 to 4 (1, no change; 2, little change; 3, moderate change; 4, large change.
SD, standard deviation.

**Table S4:**
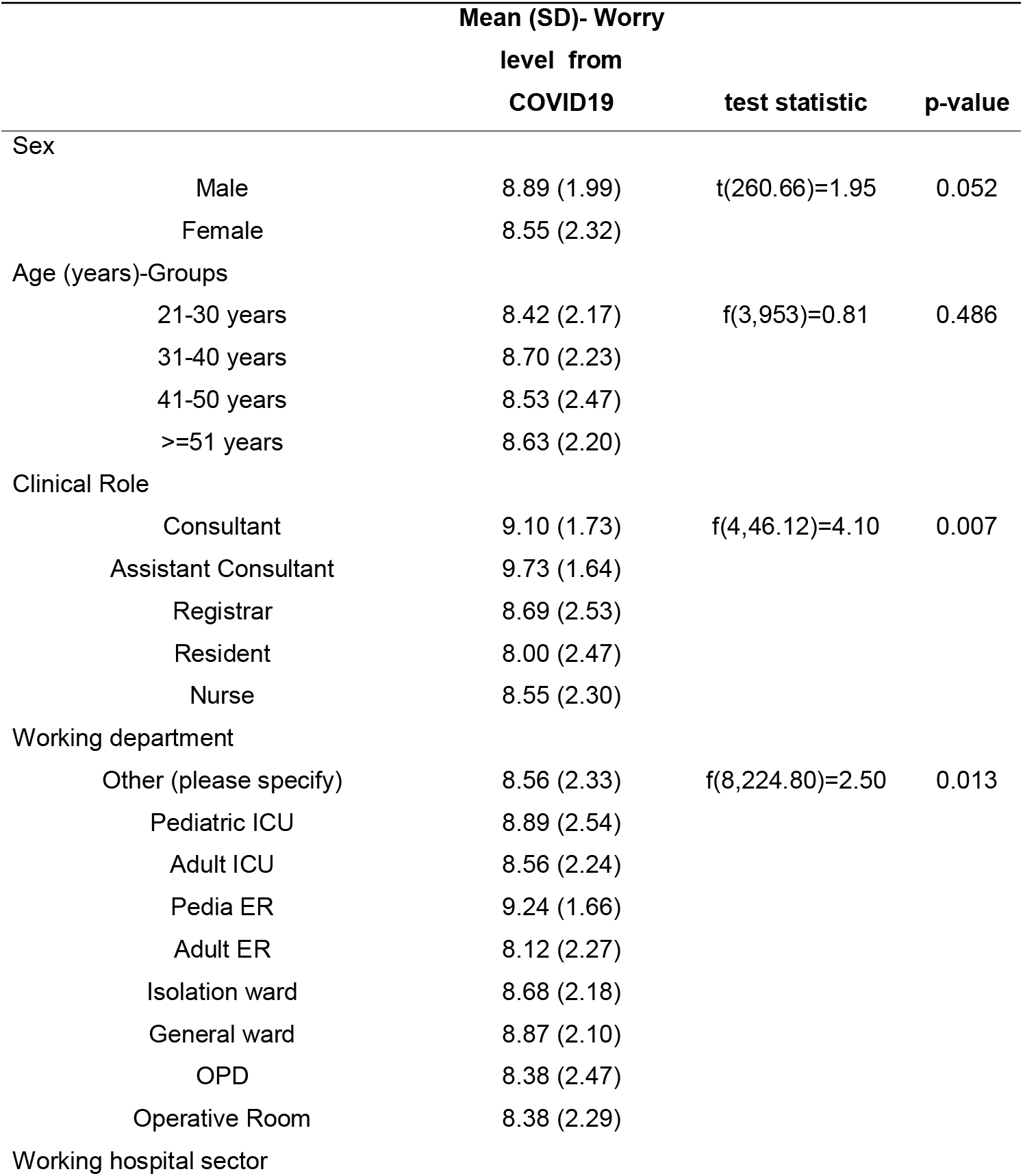
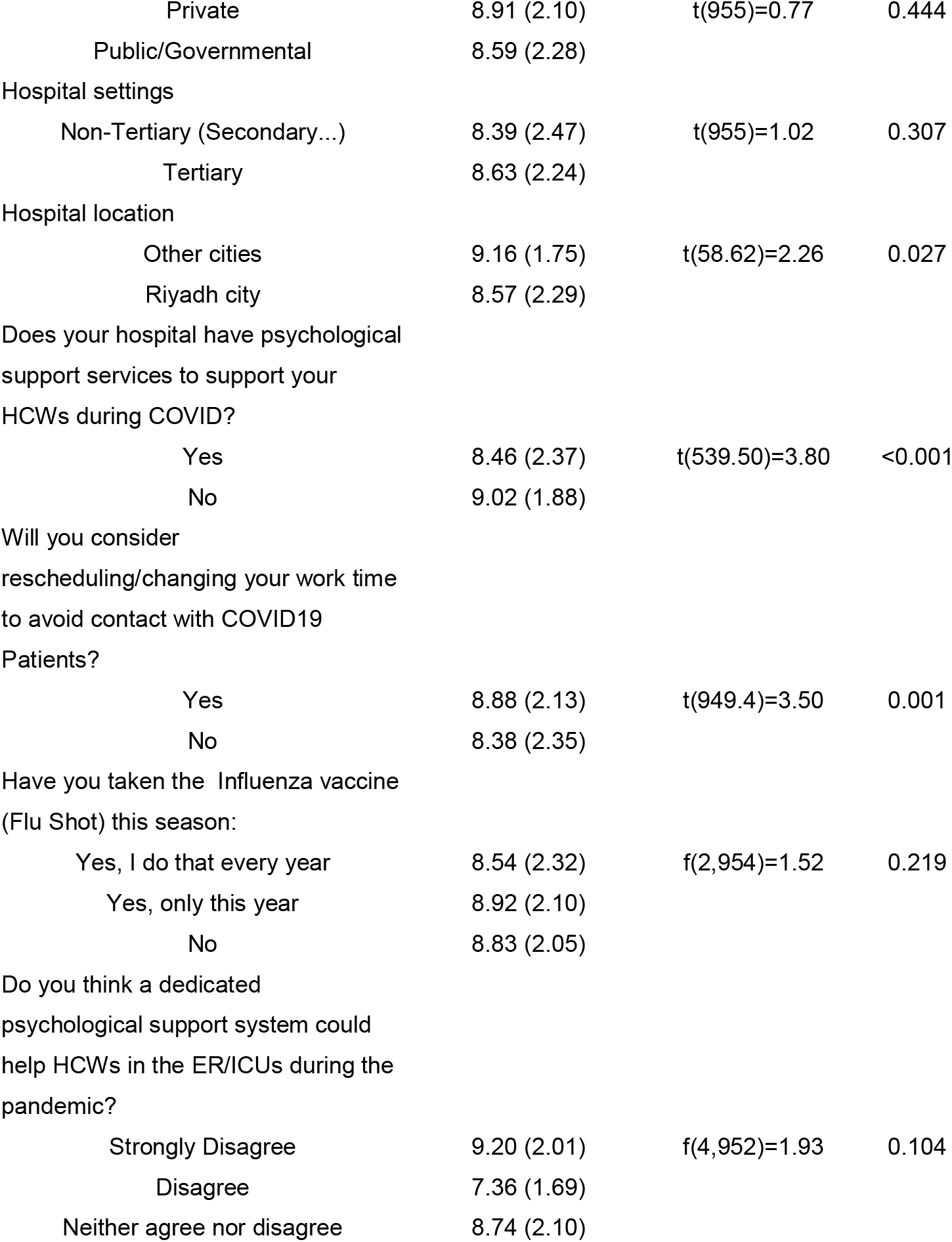
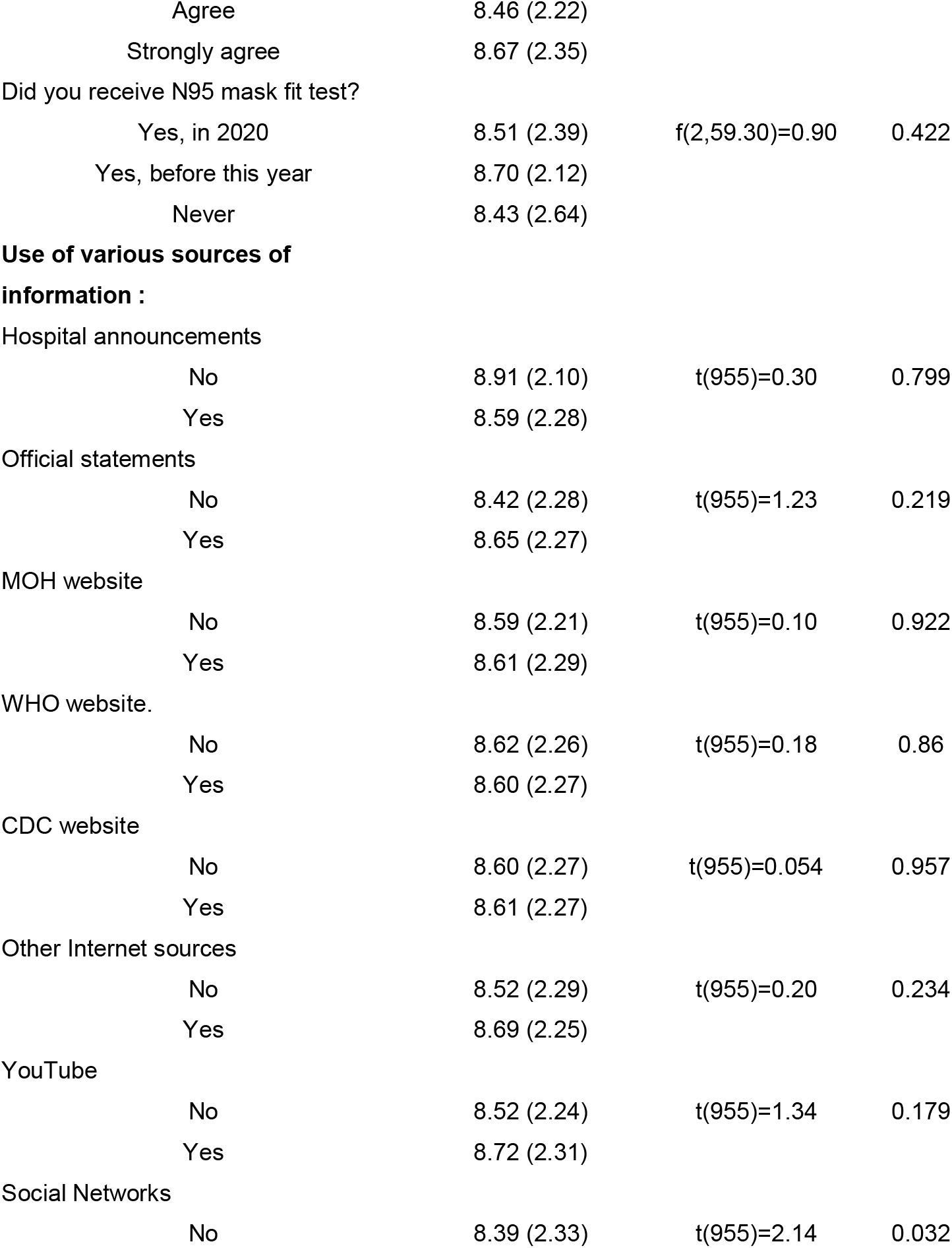
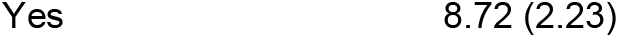
Bivariate analysis of the HCW’s current perceived Anxiety from COVID19 disease across the levels of their demographic & professional characteristics and perceptions.

## Notes

### Competing Interest Statement

The authors have declared no competing interest.

### Author Declarations

Approved by the Institutional Review Board, King Saud University, Riyadh, Saudi Arabia, Approval # 20/0064/IRB

